# A rapid CRISPR/Cas12a-based assay for the detection of HIV-1 Indian Clade-C infections

**DOI:** 10.1101/2024.11.21.24317621

**Authors:** Anjli Gaur, Harsh Bhakhri, Nitesh Mishra, Shaifali Sharma, Tanu Bansal, Mani Kalaivani, Megha Brijwal, Bimal Kumar Das, Rakesh Lodha, Subrata Sinha, Kalpana Luthra

## Abstract

Early detection of HIV-1 infection is crucial to initiate anti-retroviral therapy (ART) to suppress viremia and disease progression. Herein, we developed a CRISPR/Cas12a-based HIV-1 detection assay by optimizing components for a coupled isothermal preamplification by recombinase polymerase amplification (RPA). The HIV-1 Indian Clade-C-specific conserved pol region was targeted by crRNA designed for Clade-specific detection. The CRISPR/Cas12a cleavage of the viral cDNA input is displayed as a single visually detectable outcome due to the collateral cleavage of the ssDNA-FAM-BQ reporter, enabling the rapid detection of HIV-1. The performance of the assay was evaluated by testing sera of 41 Indian Clade C HIV-1 seropositive individuals, which included 28 HIV-1 infected infant samples, HIV-1 Indian clade C genome plasmid, viral disease control DNA/RNA samples (Influenza, RSV, Parvovirus, HPIV, CMV, and HBV), and 31 healthy donor sera samples. With 96% sensitivity and 92.65% specificity for HIV-1C detection, with fluorescence and visual readout, and a capability of detection using lateral flow dipsticks, our CRISPR/Cas12a-based HIV-1 C detection assay demonstrates the potential to be developed into a robust point-of-care molecular diagnostic test for HIV-1C. Moreover, it may serve as a potential rapid NAT alternative in detecting mother-to-child transmission (MCT) of HIV-1C in infants (<2 years of age), where rapid antibody-based serology tests are rendered ineffective due to the presence of maternal antibodies.

## Introduction

Human immunodeficiency virus (HIV-1) is an infectious entity that can lead to the development of acute immunodeficiency syndrome (AIDS) in an infected individual. According to the UNAIDS global HIV-1 statistics, 39 million people globally were living with HIV-1. In 2022, 6,30,000 people died from AIDS-related illness (1). HIV-1 Clade C, being responsible for half of the global infections, accounts for 97% of HIV-1 infections in India (2). HIV-1 infections remain to be one of the global public health issues, with an effective vaccine still elusive, lifelong adherence to antiretroviral drugs is required for the effective management of the disease (3). Hence, early detection of HIV-1 is crucial for treatment initiation, prevention of transmission, and disease progression in perinatally infected infants, children, and adults (4).

The quantitative reverse transcriptase-polymerase chain reaction (qRT-PCR) is one of the sensitive nucleic acid tests to detect HIV-1 in samples with low viral RNA copies. Its high sensitivity and specificity have been considered a “gold standard” for disease diagnostics, with several approved qRT-PCR tests for HIV-1 (5–9). However, this method is not cost-effective for HIV-1 detection in resource-limited settings. In recent decades, several isothermal amplification methods, such as loop-mediated isothermal amplification (LAMP), have been developed as attractive alternatives to conventional PCR methods because of their simplicity, rapidity, and low cost. However, deploying LAMPs as efficient POC diagnostics for clinical applications is still challenging due to complex assay design and primer set optimizations requiring a trained professional (10). For the diagnosis of HIV-1 infection in infants and children less than 18 months of age, National AIDS Control Organization (NACO, India) recommends the use of a Total Nucleic Acid PCR test on a Dried Blood spot (DBS) sample of the infant to detect viral nucleic acid for diagnosis of HIV-1 infection during infancy. Serology testing is not preferred in infants and children of this age group due to plausible interference by the transplacental passage of maternal antibodies in the infant (11–13).

With advances in CRISPR/Cas-based technology, its cost-effectiveness, high sensitivity, and specificity for its target sequence provides a platform for using the CRISPR/Cas-based systems in molecular diagnostics (14). Recently, RNA-guided CRISPR/Cas nuclease-based nucleic acid detection has shown great promise for developing next-generation molecular diagnostics technology due to its high sensitivity, specificity, and reliability. For example, some Cas nucleases (e.g., Cas12a, Cas12b, and Cas13a) perform intense collateral cleavage activities in which a crRNA-target binding activated Cas can indiscriminately cleave surrounding non-target single-stranded nucleic acids (15–18). By combining with a sensitive isothermal preamplification at 37°C using recombinase polymerase amplification (RPA) (19–21), Cas13 and Cas12a have, respectively, been used to develop SHERLOCK (Specific High-sensitivity Enzymatic Reporter UnLOCKing) system and DETECTR (DNA Endonuclease-Targeted CRISPR Trans Reporter) system for highly sensitive nucleic acid detection (22,23). Apart from the RPA method, some CRISPR-Cas-based nucleic acid sensors utilized the LAMP and PCR approaches, such as the CRISPR-Cas12b-assisted HOLMESv2 platform (24,25). The CRISPR/Cas systems, such as the CRISPR/Cas9, CRISPR/Cas12, and CRISPR/Cas13, are described for the detection of various viral nucleic acids viz, Zika virus, Ebola virus, Dengue virus, SARS-CoV2, and HBV (26–33). In the present study, we developed a sensitive CRISPR/Cas12a-based assay for rapid and specific detection of HIV-1 Indian clade C, being the most prevalent infecting HIV-1 clade in India. A crRNA targeting the conserved pol regions in the HIV-1 Indian clade C genome was used for the specific detection. The assay components were optimized for isothermal nucleic acid amplification by RPA and CRISPR/Cas12a-based cleavage reaction to occur in a single tube at 39 °C for 45 min, thereby eliminating the need for an intermediate nucleic acid purification step. The assay is adaptable for versatile detection modes, such as visual (UV transilluminator), Fluorescence intensity readout, and Lateral flow dipstick-based readout. Herein, we have developed a rapid and sensitive CRISPR/Cas12a-based HIV-1 C detection assay that demonstrates the ability to detect HIV-1 infection in infants, with the potential to be developed into a sensitive lateral flow-based point-of-care molecular diagnostic test for HIV-1 Clade-C.

## Material and methods

### Clinical samples and ethical statement

A total of 41 HIV-1 positive donor sera (including 28 infant sera) and 31 healthy donor sera, separated from blood samples that were collected from the Department of Pediatrics and Department of Microbiology, AIIMS New Delhi, India, were included in this study. All the donors were ART-naïve at the time of diagnosis and were not showing symptoms of AIDS progression. The detailed demographics and immunological profile (Table S1) were obtained from the Department of Microbiology, AIIMS New Delhi, India. Written informed consent was obtained from the parent/LAR of the infants/children. The study was reviewed and approved by the Institutional Ethics Committee (IEC) of All India Institute of Medical Sciences (IEC-114/06.03.2020), RP-47/2020. The demographic profile of the study participants was recorded systematically. DNA/RNA samples of Influenza, Respiratory syncytial virus (RSV), Parvovirus, Human Parainfluenza virus (HPIV), and Cytomegalovirus (CMV) were obtained from the Virology Testing Laboratory, Department of Microbiology, AIIMS New Delhi and used as disease controls. The HIV-1 genome plasmid 93IN999 and 93IN905(positive control) and HIV-1 Clade B infected donor sera samples (disease control) were obtained from the NIH AIDS reagent program.

### HIV-1 RNA extraction and cDNA synthesis

Viral RNA was extracted from 140 μl of serum using the QIAamp Viral RNA Mini Kit according to the manufacturer’s instructions. Immediately after RNA extraction, reverse transcription was performed using the gene-specific primer OFM19 (5’-GCACTCAAGGCAAGCTTTATTGAGGCTTA-3’) and Superscript IV reverse transcriptase (Thermo Scientific). The procedure of viral extraction to cDNA preparation using Superscript IV reverse transcriptase was performed within a one-hour duration.

### crRNA and RPA primer designing

crRNA sequence for HIV-1 clade C viruses in the pol region was designed. A total of 86 HIV-1 Clade C sequences were obtained from the HIV database (https://www.hiv.lanl.gov/content/sequence/HIV/mainpage.html). The sequences obtained were aligned on MEGAX to generate a multiple sequence alignment (MSA). The Consensus maker tool at the LANL HIV database was used to create the consensus sequence of the pol region (Figure S1). The consensus sequences within the pol region were scanned across the MSA of Indian pol using the Analyse-ALIGN tool at LANL HIV database for maximum conserved regions. Regions with maximum conservation were used as crRNA target sequences. One consideration for crRNA design for CRISPR/Cas12a-based detection assay is the lack of overlap with RPA amplicon primers, as overlap with primers may result in false positives. The identified conserved region was picked as the crRNA target, and along with a Cas12a adaptor sequence, the crRNA was synthesized from IDT (Integrated DNA Technologies). The flanking conserved regions were identified to design RPA primers with optimal conditions: size range within 30-35 and Tm range between 37℃-42℃. A primer pair was designed to amplify 183 bp RPA amplicon, which carries the crRNA target sequence RPA_Forward (5’-ACAGGRGCAGATGATACAGTATTAGAAGA-3’) and RPA_Reverse (5’-CCAATTATGTTGAYAGGTGTDGGTCC-3’).

### Probe design for fluorescence and lateral flow-based detection

The ssDNA-FAM-BQ reporter, a five-nucleotide oligonucleotide (5’-TTATT-3’) labelled by a 5’ 6-FAM (Fluorescein) fluorophore and a 3’ Iowa Black® FQ quencher-ssDNA-FAM-BQ probe (IDT) was designed and synthesized for fluorescence/visual readout. Upon the detection of the HIV-1 pol sequence by the CRISPR/Cas12a complex, the collateral cleavage activity cleaves the ssDNA-FAM-BQ, which releases the FAM molecule from the quencher, and the signal generated was detected using a UV transilluminator/lamp and fluorescence reader. The ssDNA probe with the same sequence as for fluorescence readout was used with 5’ FAM, and 3’ Biotin was designed and synthesized for lateral flow strip-based assay readout.

### The CRISPR/Cas12a-based assay reaction

The RPA reactions were performed using the TwistAmp Basic kit (TwistDx) as instructed by the manufacturer. For the CRISPR/Cas12a reaction, purified LbaCAs12a (IDT) was used. The assay was set up with RPA reagents with optimized concentrations (Figure S3 and S4):1X rehydrated Master mix, RPA primers (IDT) (168nM), and 14 mM MgOAc) and CRISPR/Cas12a cleavage reaction reagents: LbaCas12a (0.64µM), crRNA(0.64µM), 1X Cas12 activity Buffer (IDT), ssDNA-FAM-BQ fluorescent probe (8 µM) into a single tube. The reaction was incubated at 39°C for 45 min.

### Visual, fluorescence, and lateral flow dipstick-based readouts of the assay

The end-point fluorescence signal detection was done using a UV lamp/transilluminator (visual detection) and fluorescence detection by a fluorometer. The emission was recorded as AFU (Arbitrary fluorescence unit) at excitation at 488nm and emission at 520nm. For adapting the assay as a lateral flow strip-based CRISPR/Cas12a detection assay, the same reaction was set up with the ssDNA-FAM-Biotin probe with the optimized concentration (0.009 µM) such that there is no non-specific detection at the test line (Figure S6). Post incubation, 10μl the reaction mix was applied to the lateral flow dipsticks (HybriDetect - Universal Lateral Flow Assay Kit). The dipsticks were dipped into 100μl of the HybriDetect assay buffer provided with the kit. The assay buffer was allowed to adsorb for 2-3mins and colour development on the test and control line was observed to determine a positive or negative reaction.

### Statistical analysis

All the statistical analyses were performed using Python (version 3.9). The sensitivity and specificity of the diagnostic assay were calculated using the scikit-learn library. To distinguish between positive and negative HIV-1 detection using the assay, the optimal AFU threshold value for the fluorescence readout was determined by plotting the ROC curve and calculating AUC and Youden’s Index by using the ‘roc_curve’ and ‘auc’ function of the ‘scikit-learn’ library. The percentage sensitivity and specificity of the assay were computed using the ‘confusion_matrix’ function of the scikit-learn library. The Clopper-Pearson confidence intervals for specificity and sensitivity percentages were calculated using the ‘binom’ function by fitting a binomial distribution for 95% confidence. Cohen’s Kappa coefficient was also calculated to assess the agreement between the assay results and the true labels, accounting for chance agreement, using the ‘cohen_kappa_score’ function from the same library. Data plotting was performed using the GraphPad Prism software (Version 9.0; La Jolla, CA, USA).

### Data availability

All the data to support the findings of the study are available within the paper and supplementary information. Any additional information can be requested from the corresponding authors. The source data for the endpoint fluorescence readout and assay performance evaluation are provided as Source Data File of this paper.

## Results

### The CRISPR/Cas12a-based assay system

In this study, we developed and evaluated a CRISPR/Cas12a-based assay system for the rapid detection of HIV-1 Clade C viral nucleic acid in serum samples. In the assay, the preamplification step by recombinase polymerase amplification (RPA) and cleavage by CRISPR/Cas12a recognition and subsequent cleavage of ssDNA-FAM-BQ reporter to give a detectable fluorescence signal as output was standardized to work as a coupled reaction in a single tube (Figure 1a). The assay utilized a crRNA that targeted a conserved HIV-1 C pol region within the RPA amplicon (Figure 1b). The cas12a and crRNA were added into the reaction solution containing the RPA primers, recombinase, single-stranded DNA binding protein (SSB), strand– displacement DNA polymerase, single-stranded DNA fluorophore-quencher (ssDNA-FAM-BQ) reporter and target template. During the incubation of the assay reaction at 39°C for 45 minutes, the RPA amplification was initiated, and the activity of the strand displacement DNA polymerase determined the target sites for the cas12a-crRNA complex. The binding of the Cas12a-crRNA complexes with the target sites on the amplified viral DNA led to activation of the collateral cleavage activity of Cas12a endonuclease and cleavage of the ssDNA-FAM-BQ reporter molecules, generating a strong fluorescence signal. Thus, the amplified target sequences generated during the RPA triggered CRISPR/Cas12a-based collateral cleavage activity in this assay. Previous studies have demonstrated that the collateral cleavage activity of the CRISPR/Cas12a system is independent of target strand cleavage, showing that the collateral cleavage of ssDNA-FAM-BQ reporters by the Cas12a nuclease is triggered by the binding of crRNA to target sites (16,17). The fluorescence signal from the cleaved ssDNA-FAM-BQ reporter was detected using a UV transilluminator for a rapid qualitative assertion of the assay results or by taking the fluorescence readout as arbitrary fluorescence units (AFU) by a fluorescence reader at an excitation of 488nm and emission of 520nm, for a more accurate/semi-quantitative assertion of assay results and distinguish between a positive or negative CRISPR/Cas12a based cleavage reaction, hence presence or absence of HIV-1 clade C nucleic acid in the sample evaluated (Figure 1a).

**Figure 1:**
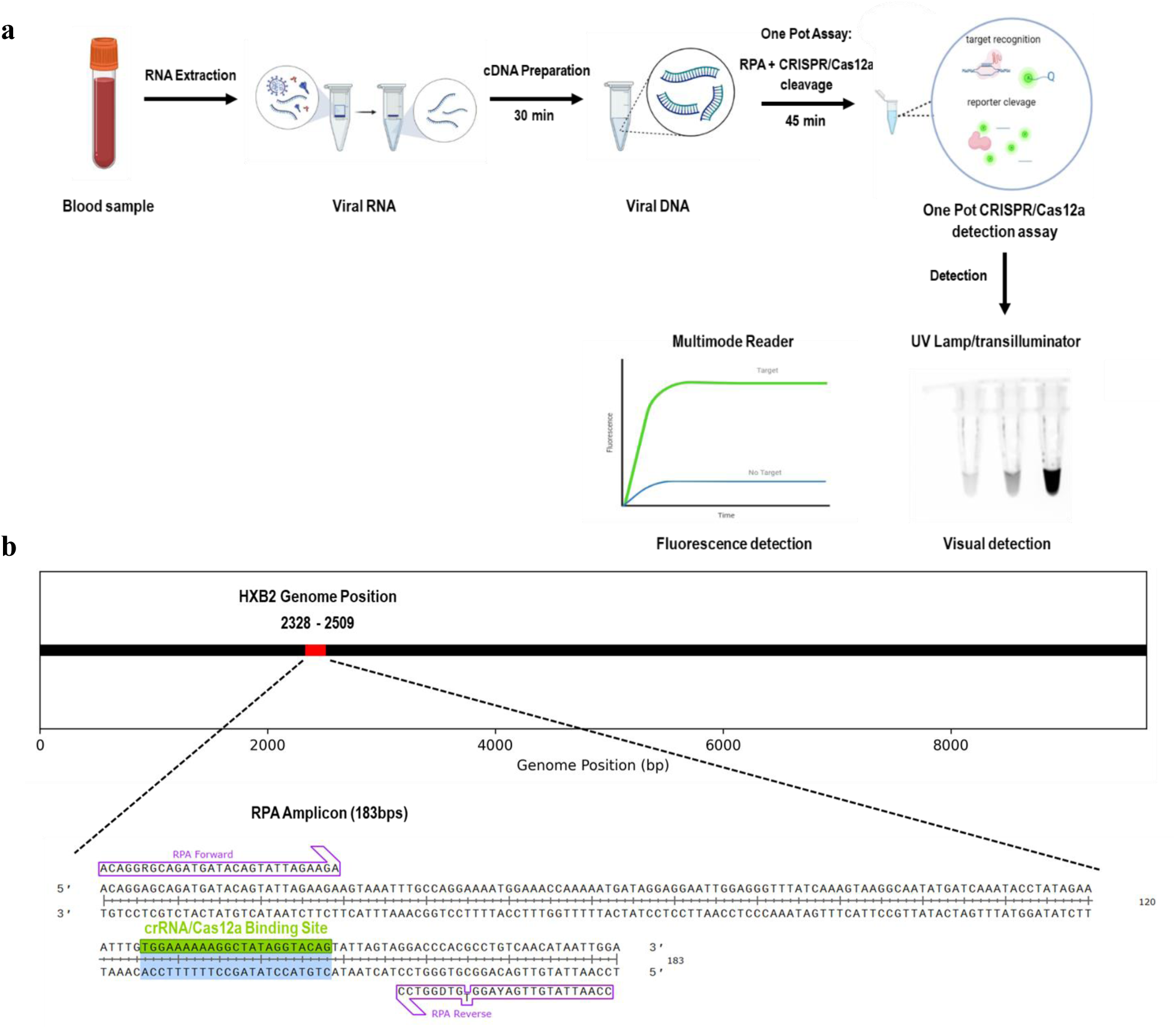
The CRISPR/Cas12a-based HIV-1 detection assay. **a)** Schematic of the CRISPR/Cas12a-based assay workflow. Viral RNA extraction followed by cDNA preparation from the serum samples yields the target nucleic acid for the assay. The RPA and CRISPR/Cas12a cleavage reaction co-occur in a single tube to aid in the detection of target nucleic acid. Cleaved reporter signal can be detected by UV lamp/transilluminator (visual detection) and multimode fluorescence reader (fluorescence detection for semi-quantitation of the assay). **b)** HIV-1 Genome map depicting the RPA amplicon region carrying the crRNA sequence. The region marked in red represents the RPA amplicon (183bps) with HXB2 coordinates 2328-2509 bps; RPA primers labelled in the purple and green region represent the crRNA sequence.

### Design and optimization of the CRISPR/Cas12a-based assay system

Various components were designed for the CRISPR/Cas12a-based detection assay to work in sync with the coupled isothermal RPA reaction, and concentrations were optimized for distinct positive vs. negative results. Firstly, the crRNA was designed to target the HIV-1 clade C sequence. The crRNA in the assay was designed to target the most conserved region of the HIV-1 Indian Clade C sequences. Upon multiple sequence alignment of 86 Indian Clade C sequences, we identified the Pol region of the HIV-1 genome as the most conserved region (Figure S1a-b). The crRNA was designed by taking 21bps of the conserved region sequence (2453-2475 HXB2 coordinate) downstream to the PAM sequence (TTTV) along with the Cas12a binding adaptor sequence (Figure S2a-b). To detect the specific region by Cas12a cleavage, preamplification by RPA was done by designing RPA primers using the conserved regions to amplify a 183bp RPA amplicon (Figure S3a), which lies in the Pol gene sequence at the location from 2328 to 2509 in the HXB2 genome map (Los Alamos). The primers were designed by keeping the range of the number of nucleotides within 30-35 and Tm in between 37℃-42℃ (Figure 1b). Further, we optimized the RPA reaction for various parameters: reaction time, primer concentration, and sample cDNA amount. Multiple reactions were set up to determine the optimal time for RPA by incubation at different time points from 20 minutes to 40 minutes, with an increment of 5 minutes. An incubation of 30-40 mins was optimal for amplifying the HIV-1 amplicon (Figure S3b). RPA reactions with a gradient of primer concentrations were performed to determine the optimal primer concentration. Optimal amplification was observed at a primer concentration of 168nM (Figure S2c). To determine the patient’s cDNA dilution to perform RPA, an HIV-infected infant donor cDNA sample of AIIMS748 was tested from neat to 1:20 cDNA dilutions. Low amplicon band intensity was observed at neat and 1:5 cDNA dilution, with a darker band intensity at 1:10 and 1:20 cDNA dilution depicting hindrance of cDNA component at lower dilution with optimal detection at a higher cDNA dilution (Figure S3d). The CRISPR/Cas12a-based cleavage assay was optimized for the detection of HIV-1 in clinical samples. The ssDNA-FAM-BQ reporter concentration (8 μM) and the ratio of crRNA to cas12a (0.64nM) (1:1) were used for the CRISPR/Cas12a-based cleavage reaction (29). For a coupled RPA and CRISPR/Cas12a-based cleavage for detection of HIV-1 target sequence, all the above-optimized assay components were incubated at 39°C for 45 minutes for all the amplified sequences to be cleaved simultaneously. The CRISPR/Cas12a-based HIV-1 detection assay was performed for HIV-1 clade C genome plasmids 93IN999 and 93IN905 (10 ng each), and multiple cDNA dilutions from one HIV-1 positive infant donor AIIMS748 were used for the optimization of the assay. Corroborating with the RPA amplification of patient viral cDNA, a cDNA dilution of 1:10 yielded a better detectable fluorescence and visual readout signal (UV transilluminator) (Figure 2a-b). Hence, all the HIV-positive patients’ viral cDNA samples were tested at a minimum of 1:10 cDNA dilution.

**Figure 2:**
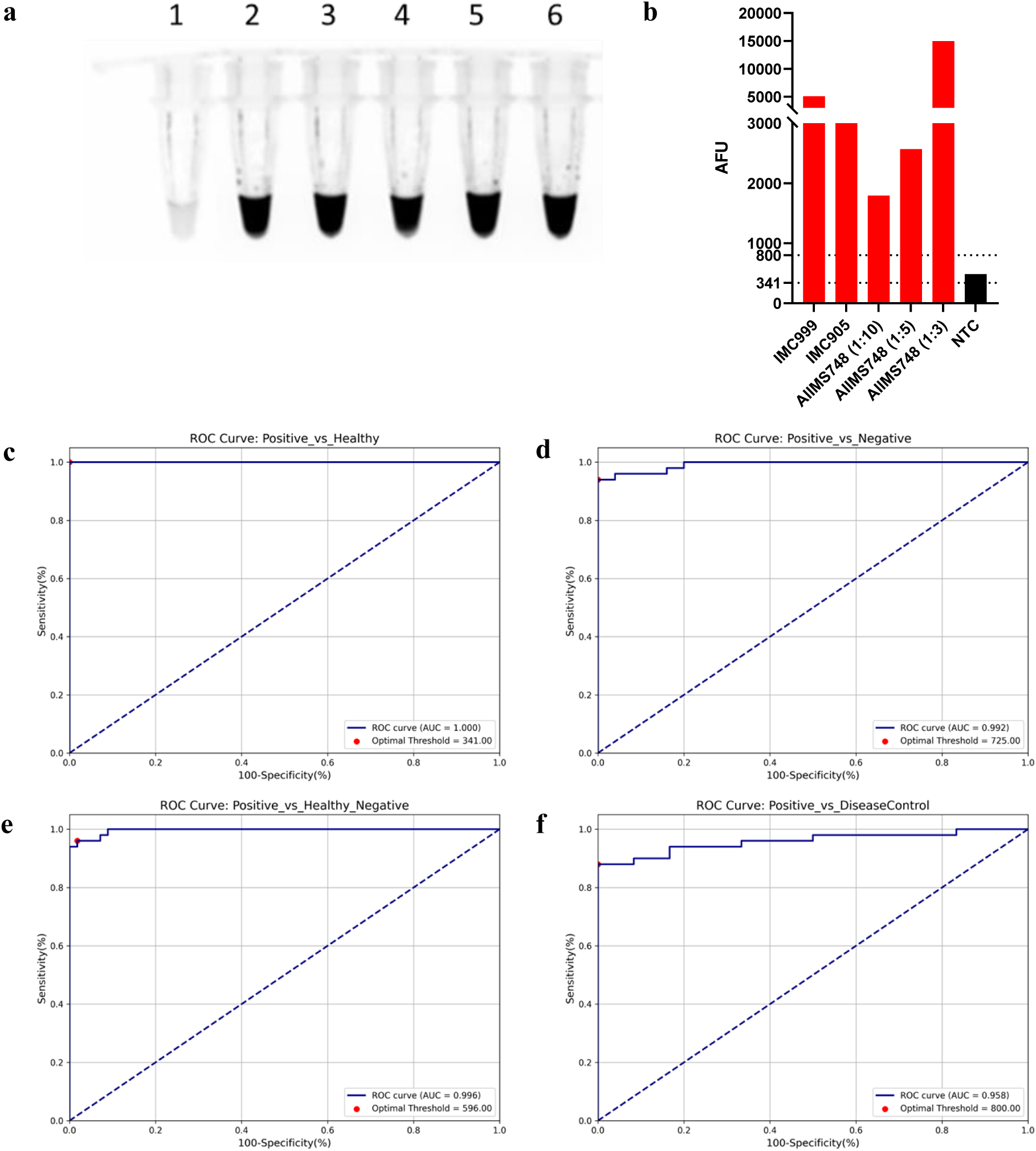
Evaluation of the CRISPR/Cas12a-based assay for detecting the HIV-1 Indian Clade C in clinical samples. The CRISPR/Cas12a-based HIV-1 clade C detection assay was validated using positive controls. **a)** UV transilluminator image and **b)** fluorescence readout of the assay performed for the AIIMS748 (HIV-1 positive donor) cDNA at different dilutions and in the HIV-1 genome plasmids (93IN999 and 93IN905). Lane 1-NTC, lane 2-93IN999, lane 3 - 93IN905, lane 4-AIIMS748(1:10) dilution, lane 5-AIIMS748(1:5) dilution, lane 6-AIIMS748(1:3) dilution. The CRISPR/Cas12a-based assay results were recorded using visual (UV transilluminator) and fluorescence (AFU) readout. The fluorometer readings of fluorescence emission were recorded as AFU (Arbitrary fluorescence unit) at excitation at 488nm and emission at 520nm. **c-f)** The ROC curves are plotted for the evaluation of the assay performance and to determine optimal AFU threshold values for distinguishing between an HIV-positive and negative sample. The ROC analysis was done between clinical samples groups; Positive vs Healthy (AUC-1.000), Positive vs Negative (AUC-0.992), Positive vs Healthy and Negative (0.996), Positive vs DiseaseControl (AUC-0.958) to determine the optimal threshold by Youden’s index as 341, 725, 596 and 800 respectively. The optimal AFU obtained from ROC curves from the respective groups were used to evaluate the sensitivity and specificity of the assay.

### Evaluation of sensitivity and specificity of the CRISPR/Cas12a-based HIV-1 C detection assay in clinical samples

The high sensitivity and specificity of an assay are a testament to its effectiveness. We evaluated the sensitivity and specificity of our assay using ‘True Positives’ and ‘True Negatives’ as test samples. True positives comprised of the cDNA samples from 41 HIV-1 positive donor sera and 9 replicate samples of HIV-1 genome plasmid 93IN999 and 93IN905. True negatives comprised of cDNA samples from 31 healthy donor sera, viral DNA/RNA samples of 15 viral disease controls (1-Influenza, 1-RSV, 1-Parvovirus, 1-HPIV, 3-CMV, and 5-HBV genome plasmids), and 25 non-template control (NTCs) replicates (Figure 3a). Firstly, the Receiver operating characteristic (ROC) curves using the arbitrary fluorescence unit (AFU) values for the various samples tested were plotted to distinguish between the positive and negative results of the assay and determine the optimal AFU threshold values. The ROC curves were plotted separately for positive and different negative sample groups: positive vs healthy controls, positive vs negative controls (NTCs), positive vs healthy and negative controls, and positive vs disease controls. The area under the curve (AUC) and Youden’s Index were calculated for each ROC curve to determine the optimal AFU threshold values of fluorescence readout above which any sample tested can be classified as positive for the assay. Optimal AFU threshold values to determine test positivity in the context of the different controls; 341 (positive vs healthy-AUC 1.00), 725 (positive vs NTCs-AUC 0.992), 596 (positive vs healthy and NTCs-AUC 0.996) and 800 (positive vs disease controls-AUC 0.958) (Figure 2c-f). Since we observed an overlap between some of the positive samples with the lowest AFU and that of the highest AFU of a few negative samples, we defined the sensitivity and specificity of the assay by taking into account all the different AFU threshold values obtained. The AFU threshold of 725 yielded a maximum sensitivity and specificity of 94% and 97.03%, respectively, that led to 3 false negatives (FN) and 2 false positives (FP) (Figure 3d). The AFU threshold of 596 yielded 96% sensitivity and 92.65% specificity, with 2 FNs and 5 FPs (Figure 3e). The lowest and highest AFU threshold values, 341 and 800, result in compromised specificity and sensitivity, respectively (Figure 3c, 3f). Using the AFU threshold of 341 yielded 0 FNs but 15 FPs, mostly from viral disease controls and NTCs values, suggesting that this threshold value could serve as a good baseline for screening for HIV-1C in a population. Also, AFU threshold of 800 yielded 0 FPs but 6 FNs, and could serve as an ideal AFU threshold, as any value above this threshold may confirm the presence of HIV-1C nucleic acid. Based on these observations, we defined a range of AFU threshold of 341-800, with AFU values of test samples <341 and >800 would be described as negative and positive for the assay respectively, while samples with AFU values between 341-800 would be reported as inconclusive, with a need for reassessment and validation (Figure 3b).

**Figure 3:**
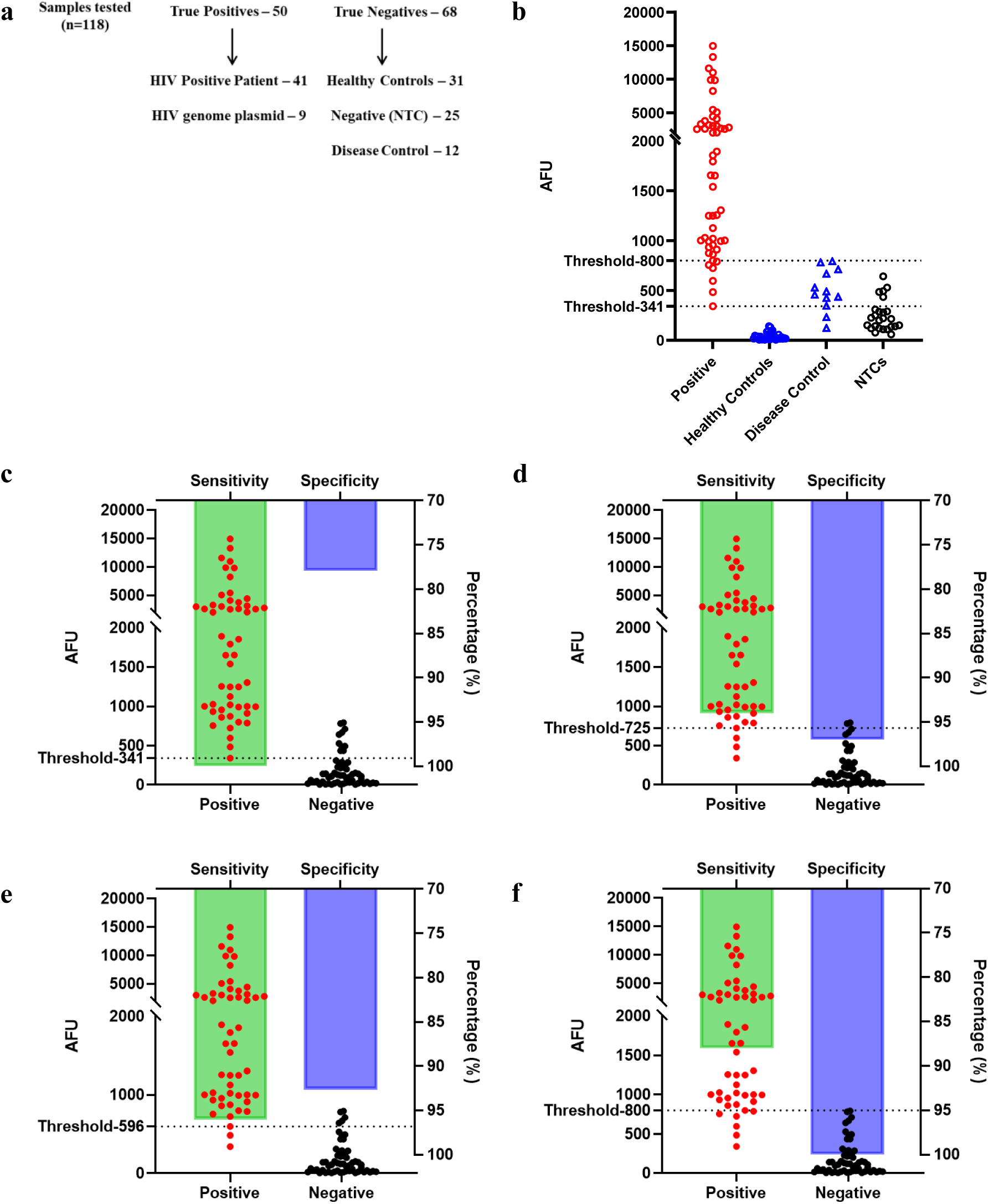
Performance of the CRISPR/Cas12a-based assay in clinical samples. **a)** Schematic of assay performance evaluation using true positives-50 (41 HIV positive plasma, 9 HIV-1 genome plasmid 93IN999 replicates) and true negatives-68 (25 NTC replicates, 31 healthy donor plasma samples, 12 viral disease control DNA/RNA samples), total-118 samples, to plot an ROC curve and determine the optimal AFU threshold and evaluate the sensitivity and specificity of the assay in detecting HIV-1C in clinical samples. **b)** Comparison of the AFU values for the different clinical samples tested for the presence of HIV-1 using the CRISPR/Cas12a-based detection assay. AFU threshold values 800 (highest) and 341(lowest), determined using the ROC curve, were used to distinguish the positive and negative results of the assay. **c-f)** The sensitivity and specificity of the assay were evaluated to determine the positive and negative samples efficiently using the different AFU threshold values obtained from the ROC curve analysis. AFU threshold values of 341, 725, 596, and 800 were used to evaluate the sensitivity and specificity of the assay, respectively. The positive group includes HIV-1 positive clinical samples and positive genome plasmid replicates, and the negative group consists of the healthy controls, NTCs, and viral disease controls. Dots represent the AFU values plotted against the left y-axis, and Bars represent the sensitivity or specificity values plotted against the right y-axis.

To evaluate the assay performance and determine the sensitivity and specificity, an AFU threshold of 596 was selected, as it yielded the least FNs with high specificity and sensitivity values of 96% and 92.65%, respectively, and a significant kappa value of 0.879 (Table 1). The viral cDNA samples of 41 HIV-1 positive sera from clade C-infected infants/children and adults were tested in the assay. Of the 41 samples, 22/41 were of known viral load, determined by qRT-PCR, and 19/41 were seropositive for HIV-1 IgG/IgM (with viral load not determined), which were included to broaden the evaluation of the assay (Table S1). 35/41 positive clinical samples were tested positive, and 6/41 positive samples were reported as inconclusive for the assay using the AFU threshold range established (Figure S4a). Our findings demonstrated that the HIV-1C detection assay developed herein possesses high sensitivity and specificity, affirming its effectiveness. The CRISPR/Cas12a-based assay tested positive for the lowest viral load sample with 117000 copies/ml at a cDNA dilution of 1:10 from the donor AIIMS744 (Figure S4a, Table S1). Of the 41 cDNA samples of HIV-1 seropositive individuals tested, 28 were from infants (<2 years) (Table S1). With 27/28 HIV-1 positive infant samples tested positive using this assay and only 1/28 as result inconclusive, the assay yielded a sensitivity of 96.43%, demonstrating the ability to effectively detect HIV-1 in infant sera samples (Figure S4b). Serology-based methods are not suitable for testing HIV-1 in infant samples due to the presence of of maternal antibodies. No visual or fluorescence signal for the assay was observed in 31/31 cDNA samples from healthy controls demonstrating high specificity (Figure 4c). Furthermore, the specificity of the assay was validated using viral disease control DNA/RNA samples from Influenza, Respiratory syncytial virus (RSV), Parvovirus, Human Parainfluenza virus (HPIV), Cytomegalovirus (CMV), and Hepatitis B virus (HBV). While the UV transilluminator did not show a significant signal, the fluorescence readout showed an AFU value in the range of 341-800 for most of the samples tested, hence falling in result inconclusive criteria (Figure 4a). This depicts that with the presence of other viral nucleic acid, there is an increase in the background fluorescence. Hence, while distinguishing between HIV-1C and other viral diseases, samples with tested AFU value between 341-800 should be retested by RT-PCR prior to reporting for HIV-1 and other viral diseases. Further, we evaluated the assay for the ability to discriminate HIV-1 Indian clade C from clade B HIV-1. The assay showed specific detection of HIV-1 clade C in positive plasmid and HIV-1 positive donor cDNA sample AIIMS748, with no detection in 7 different clade B donor sera (Figure 4b). A positive signal for HIV-1 clade C and no signal for HIV-1 clade B sequences demonstrates that the assay specifically detects the major infecting HIV-1 Clade in India. Thus, our CRISPR-based HIV-1 Clade C detection assay offers a simple, rapid, and visual approach to HIV-1 Clade C detection.

**Figure 4:**
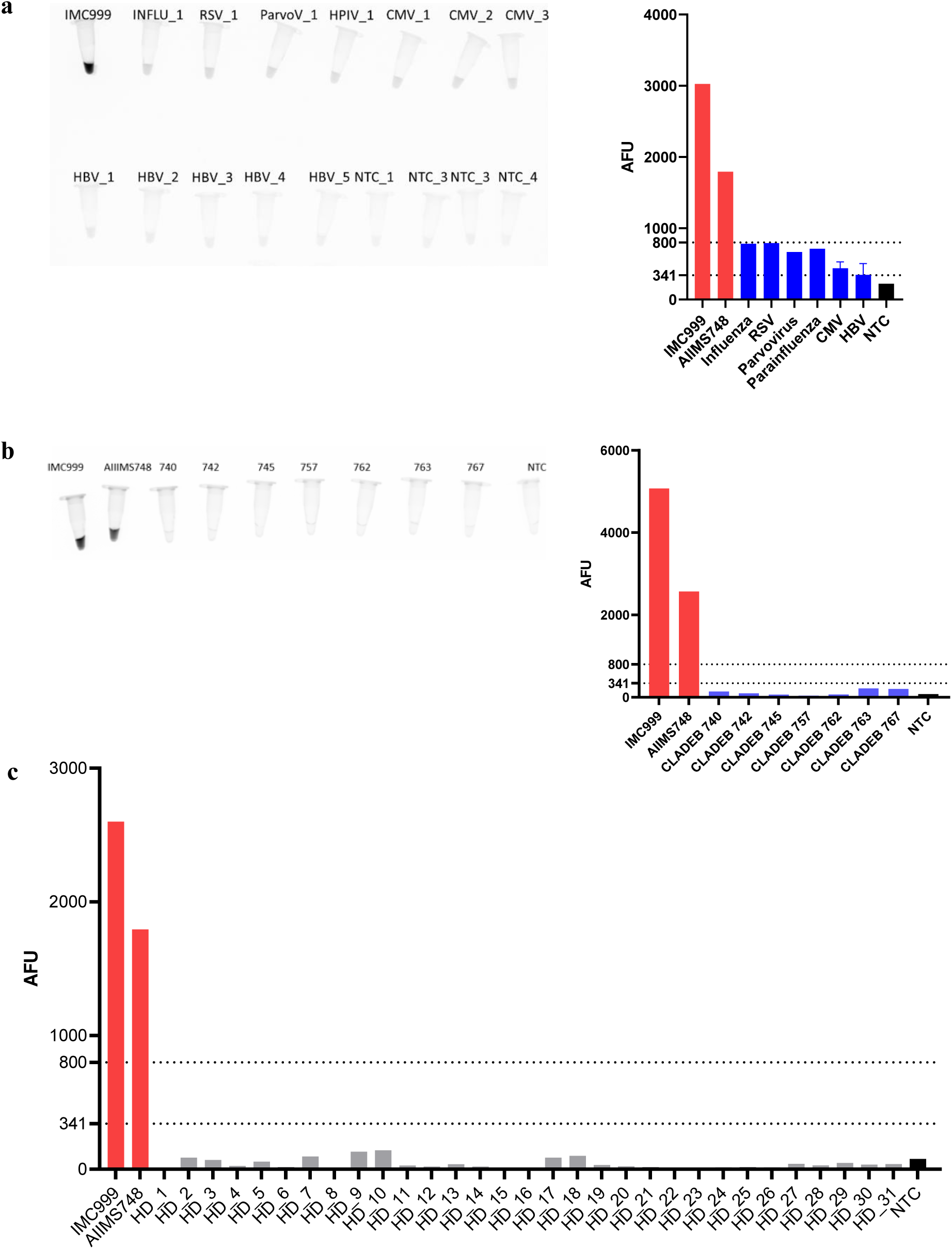
Specificity of the CRISPR/Cas12a-based assay for detecting HIV-1 Indian clade. **C.** The specificity of the assay was evaluated against different HIV-1 negative biological samples, and the output in the form of UV transilluminator (visual) readout and fluorescence detector readout AFU (Arbitrary fluorescence unit) was compared across all samples. **a)** Evaluation of assay specificity using viral disease control samples n=15; Influenza virus RNA sample from patient plasma, Respiratory syncytial virus (RSV) RNA sample from patient plasma, Parvovirus positive DNA sample from patient plasma, Human Parainfluenza virus (HPIV) RNA sample from patient plasma, CMV positive DNA sample from patient plasma and HBV genome plasmid replicates. **b)** Evaluation of clade specificity of the detection assay in distinguishing HIV-1 Indian clade C infections using n=7 HIV-1 clade B serum samples. **c)** Evaluation of assay specificity using n=31 Healthy donor serum samples. The assay specificity was validated by using positive controls (HIV-1 Indian clade C genome carrying plasmid 93IN999 and serum sample of Indian clade C infected patient AIIMS748) for HIV-1 Indian clade C detection along with the different negative controls tested. The fluorometer readings of fluorescence emission were recorded as AFU (Arbitrary fluorescence unit) at excitation at 488nm and emission at 520nm. AFU threshold values 800 (highest) and 341(lowest) were used to distinguish positive and negative results of the assay.

**Table 1:** Performance characteristics of the CRISPR/Cas12a-based HIV-1 C detection assay.

| CRISPR Cas12a-based assay results<br>(Fluorescence readout) | ROC Curve |  | Confusion Matrix (All Samples) |  |  | Performance Metric |  |  |
| --- | --- | --- | --- | --- | --- | --- | --- | --- |
|  | AUC | Optimal AFU Threshold<br>(Youden's Index) | Tested\True | qRT-PCR/Serology |  | Sensitivity<br>(%, 95% CI) | Specificity<br>(%, 95% CI) | Kappa Value |
|  |  |  |  | Positive | Negative |  |  |  |
| Positive vs Healthy Controls | 1.000 | 341.00 | Positive | 50 | 0 | 100.00<br>(100.00-100.00) | 77.94<br>(67.65-86.76) | 0.75 |
|  |  |  | Negative | 15 | 53 |  |  |  |
| Positive vs Negative Controls (NTCs) | 0.992 | 725.00 | Positive | 47 | 3 | 94.00<br>(86.00-100.00) | 97.03<br>(92.65-100.00) | 0.913 |
|  |  |  | Negative | 2 | 66 |  |  |  |
| Positive vs Healthy + Negative Controls | 0.996 | 596.00 | Positive | 48 | 2 | 96.00<br>(90.00-100.00) | 92.65<br>(85.29-98.53) | 0.879 |
|  |  |  | Negative | 5 | 63 |  |  |  |
| Positive vs Disease Controls | 0.958 | 800.00 | Positive | 44 | 6 | 88<br>(78.00-96.00) | 100.00<br>(100.00-100.00) | 0.894 |
|  |  |  | Negative | 0 | 68 |  |  |  |

### Adaptation of the CRISPR/Cas12a-based HIV-1 clade C detection assay as Lateral Flow Assay (LFA) for point-of-care diagnosis

The potential for point-of-care diagnostics is a promising aspect of our CRISPR/Cas12a-based assay. Hence, the assay was adapted for the lateral flow dipstick-based detection of the cleaved probe molecules. The ssDNA-FAM-Biotin probe has a 5’FAM molecule and 3’Biotin molecule; all the probe molecules are captured at the control line via streptavidin-biotin interaction if there is no collateral cleavage of the probe, while upon collateral cleavage of the probe in the presence of target sequence, the FAM molecule is released and captured by the rabbit anti-FAM Ab gold nanoparticles and sequestered on the test line via anti-rabbit antibodies leading to colour development (Figure 5a). Firstly, the ssDNA-FAM-Biotin probe concentration was optimized to prevent any spillage of the probe onto the test line for a false positive detection. Using negative control reactions (non-template controls (NTCs)), the concentration of the ssDNA-FAM-Biotin probe was optimized using various dilutions, initially with a broad range of concentrations from 1.00μM to 0.00005μM. The concentration range of 0.0124 M to 0.0042 M showed the least non-specific detection at the test line (Figure S5a). A narrow range probe concentration between 0.0124 μM to 0.0042 μM was tested, and 0.0124M to 0.008 M showed the least non-specific test line detection (Figure S5b). Furthermore, the non-specific detection was minimized by adding 5% PEG6000 (based on the manufacturer’s suggestions) (Figure S5c). The HIV-1 genome plasmid (93IN999) was detected using probe concentrations of 0.0124 μM and 0.009 μM with 5% PEG6000. The most optimal detection of the cleaved probe on the test line in a positive reaction with minimal nonspecific detection in the NTC reaction was observed using 0.009 μM ssDNA-FAM-Biotin (Figure 5b, S5d). Overall, our study suggests the potential of the CRISPR/Ca12a-based HIV-1 clade C detection assay to be developed into point-of-care diagnostics of HIV-1, instilling optimism about its future application.

**Figure 5:**
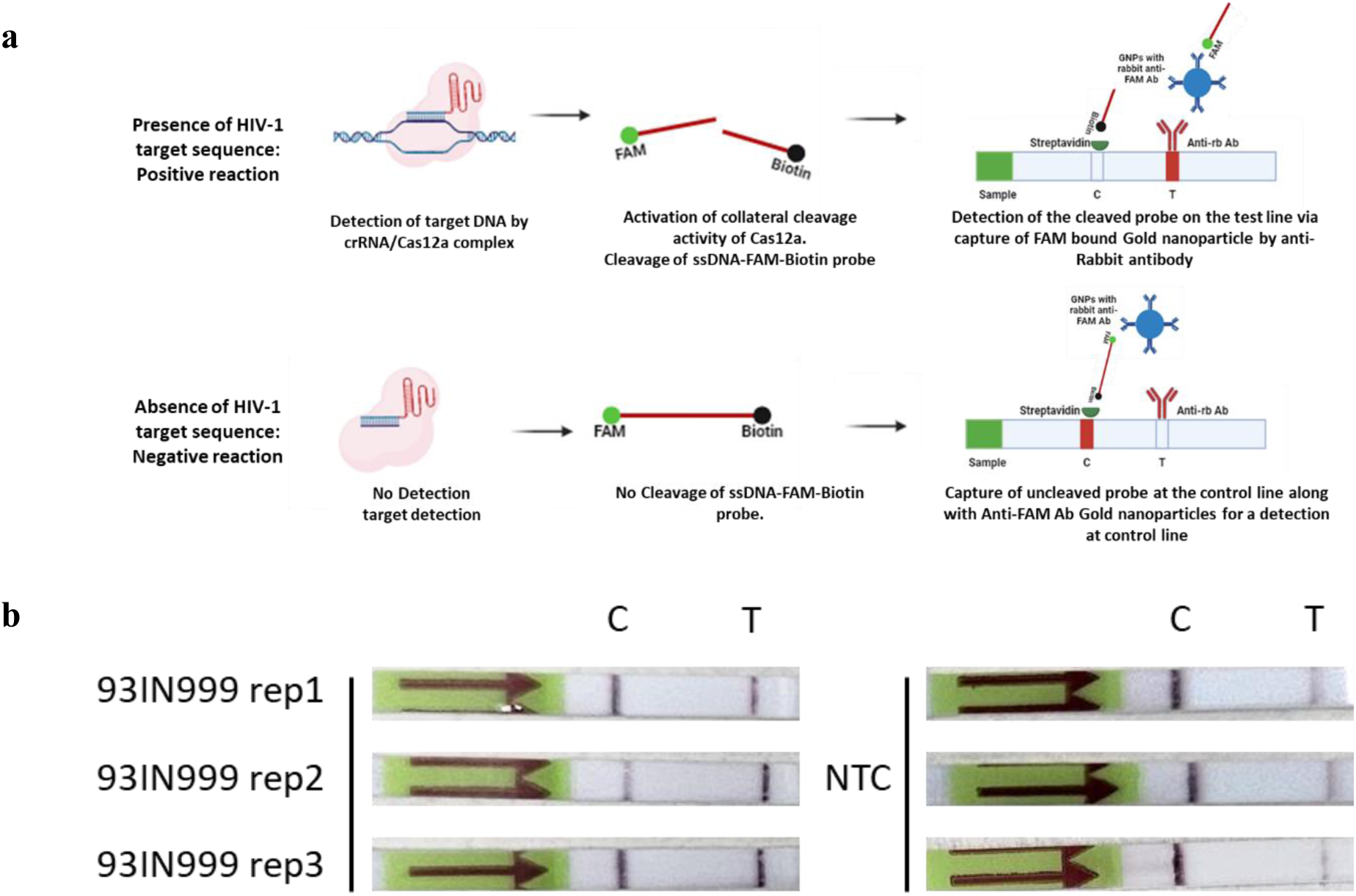
The lateral flow CRISPR/Cas12a-based HIV-1C detection assay. **a)** Schematic of Lateral flow strip-based detection of the CRISPR/Cas12a assay. ssDNA-FAM-Biotin reporter, upon activation of collateral cleavage activity of Cas12a in the presence of HIV-1 target sequence, releases the FAM molecule, which is bound by anti-FAM Gold nanoparticles and is captured by anti-rabbit antibody on the test line. In the absence of an HIV-1 target sequence, the uncleaved probe gets captured at the control line via streptavidin-biotin interaction. **b)** Detection of HIV by the CRISPR/Cas12a-based detection assay using lateral flow strips to optimize the assay into a point-of-care CRISPR/Cas12a-based lateral flow assay (LFA). HIV genome plasmid 93IN999 was used to optimize lateral flow strip-based readout of the assay with 0.009μM ssDNA-FAM-biotin probe concentration. A dark band on the test line represents a positive detection with no detection on the test line in the corresponding NTC strip.

## Discussion

Early detection of viral infections aids in the effective management of the disease by allowing timely interventions and preventing the spread. Major clinical laboratory tests for diagnosis of viral infection are based on principles of immune assays, detection of viral antigens or specific antibodies in the test sample or on the principles of nucleic acid amplification and detection of specific viral nucleic acid. Serology immune assays are rapid, point-of-care, and cost-effective tests but are not highly specific and sensitive; hence, they majorly serve the purpose of rapid screening (34). Whereas Nucleic acid amplification tests (NAAT) such as PCR, quantitative PCR (qPCR), reverse transcriptase PCR (RT-PCR) and reverse transcriptional loop-mediated isothermal amplification (RT-LAMP) are robust, highly specific, and sensitive. Hence, they are reliable confirmatory diagnostic methods but are costly and require sophisticated equipment (thermal cycler) and a trained technician, which is not practical in low-resource settings (10,35,36).

HIV-1 infections are one of the major global health concerns, with no known cure or vaccine and lifetime adherence to anti-retroviral therapy being the only effective management strategy. The diagnostic strategies for HIV-1 involve the use of serum immune assays for rapid screening of HIV-1 antigen or host antibodies, which is further confirmed by a qualitative PCR and viral load determination by qPCR for early initiation of ART and prevention of disease progression (5,8,37). Although the 4^th^ generation antibody and p24 antigen or combination laboratory immunoassays are highly sensitive and evaluated and widely deployed, but in cases of acute HIV-1 infections, antibody-based immunoassays may not be effective as seroconversion in case of HIV-1 may take 2-3 weeks. Although the patients show detectable levels of HIV-1 RNA and p24 during this period, they remain negative for immunoassays. This leads to erroneous detections during this period as the patients may have a high viral load, which leads to an increased risk of transmission (38–40). Hence, NAAT-based laboratory diagnosis plays an important role in the detection of acute HIV in combination with p24 and immunoassays. While these highly sensitive immunoassays and NAATs for HIV-1 require a sophisticated laboratory setup, with costly equipment and trained technicians, the surveillance and prevention of HIV-1 becomes less efficacious in low and middle-income countries. There are emergence of various FDA-approved point-of-care rapid HIV-1 screening tests, which detect anti-HIV IgG and IgM but fail in detecting acute HIV-1, with a 100% failure rate in case of NAAT positive/seronegative samples (41). Hence, developing a rapid point-of-care assay for HIV-1 NAAT assay, which is specific, sensitive, and cost-effective, may aid in robust HIV-1 detections and decrease the risk of spread.

Moreover, mother-to-child transmission (MCT) is even more difficult to diagnose efficiently. It is reported that the occurrence of bNAbs is more frequent in infants (>40%) (42,43) and children (44) as compared to adults, where bNAbs development occurs in a rare subset of chronically infected individuals. A potent protective antibody response in infants is governed by multivariant infections in perinatally acquired HIV-1 (42), with an improvement in bNAb response upon initiation of combined antiretroviral therapy (cART) (45). Hence, early detection of MCT is essential for the initiation of cART in perinatal HIV-1 infections. Standard serology tests such as ELISA and Western blot are not suitable for detecting HIV-1 infection in infants (below the age of 18 months), as the circulating maternal antibodies could lead to erroneous interpretation.

In this study, we have developed a CRISPR/Cas12a-based assay for rapid, sensitive, and specific detection of HIV-1 clade C. Herein, all assay components are optimized for setting up the preamplification by isothermal RPA and CRISPR/Cas12a based cleavage reaction in a single tube at 39°C for 45 mins, without the need of any intermediate purification or extraction step, thus enabling robust molecular detection of HIV-1. The UV visual readout, fluorescence-based readout, and adaptability into a lateral flow dipstick-based detection assay demonstrate the versatility and employability of the CRISPR/Cas12a-based HIV-1 detection assay developed as a potential point-of-care diagnostic test. High mutagenesis in HIV-1 is attributed to high genetic variation, which makes it difficult to target using the CRISPR/Cas12a system. Conserved regions (pol) in the HIV-1 genome were identified by aligning 86 HIV-1 sequences from Indian clade C infections. A crRNA targeting a highly conserved region in the Pol gene of the HIV-1 genome was designed and utilized in the assay for efficient detection of HIV-1. The assay was evaluated for its ability to distinguish the presence or absence of HIV-1 nucleic acid by testing various HIV-1 positive (patient cDNA and HIV genome plasmids) and HIV-1 negative samples (healthy controls, viral disease controls, and NTCs) and AFU threshold range was established using ROC curve. A range of AFU threshold value between 341-800 was used to distinguish positives from negatives efficiently, with any AFU values of the test samples less than 341 and greater than 800 determined as negative or positive for the assay, respectively, and AFU values of the test samples in between 341-800 are classified as result inconclusive. Samples with AFU readouts between 341-800 should be reassessed using RT-PCR. Evaluating the assay performance with an optimal AFU threshold of 596 with an ROC AUC of 0.996 yielded 96% sensitivity and 92.65% specificity, implying that the assay is efficient in the detection of HIV-1C in the tested samples. However, on testing disease control samples (of Influenza, RSV, Parvovirus, Parainfluenza, CMV, and HBV), our assay yielded AFU values between 341-800, suggesting an increase in nonspecific background fluorescence. This implies that test samples (including those with viral infection other than HIV-1) with a tested AFU value between 341-800 should be validated by RT-PCR to confirm the presence of HIV-1C and or other viral nucleic acid. Further, our assay specifically detects HIV-1 Indian Clade-C and not Clade B, which is the other major infecting clade of HIV-1 worldwide, hence shows that clade-specific crRNA could be designed for HIV-1, which may aid in screening for HIV-1 clade distinction in a geographical location.

With 96.43% sensitivity (27/28) in the detection of HIV-1 in infants, the assay may serve as a potential alternative to the serology tests, which are inefficient to test perinatal HIV-1 infection. Although the whole nucleic acid PCR/qRT-PCR in dried blood spots (DBS) from heel prick is one of the widely employed methods for HIV-1 detection in infants, it is less sensitive as compared to direct sera detection and requires a 2-3 hours nucleic acid extraction procedure followed by 2-3 hours of qRT-PCR reaction (46). Hence, a high turnaround time and difficulty in extracting nucleic acid from DBS leads to false positives and may require multiple sampling for validation of test in infants. On the contrary, our assay requires 200-300μl of whole blood sample for viral RNA extraction and cDNA preparation. The steps are the same as those in the DBS-qRT-PCR method, of 1 hour duration. The cDNA extracted can be directly used to detect HIV-1 in the test samples using the assay developed with a turnaround time of 45 minutes, hence involving fewer steps and time.

Similar CRISPR/Cas12a-based HIV-1 detection assays have been developed for the specific detection of Chinese clade B and circulating recombinant forms (CRFs) (47), that target a distantly conserved pol region compared to the Indian clade C pol gene conserved sequence used in our assay. There are a few limitations of our assay, such as the viral RNA needs to be prior converted to cDNA as additional steps before the assay, since the template for the CRISPR-Cas12a based assay reaction is cDNA and the limit of detection of the assay remains to be determined in terms of least number of detectable HIV-1 copies in the clinical test samples. There is scope to further simplify the steps in the present CRISPR/Cas12a-based HIV-1 detection assay and overcome the requirement of an additional step of viral cDNA synthesis by optimizing an all-in-one RT CRISPR-Cas12a assay using MMLV reverse transcriptase, that can operate at 37-40°C (hence compatible to the assay conditions), to directly detect HIV-1 RNA targets using sera as starting material. Thus, such CRISPR/Cas12a-based detection assay systems demonstrate the potential to be developed into true, handheld, point-of-care nucleic acid tests.

In summary, we have developed a robust CRISPR/Cas12a-based detection assay for clade C HIV-1. Further testing of a large number of clinical samples using this lateral flow-based detection method is warranted to evaluate the potential of the assay for point-of-care HIV-1 C molecular diagnostics in resource-limited settings.

## Acknowledgments

We are thankful to all the study subjects who participated in this study. We thank the NIH AIDS Reagent program for providing HIV-genome-carrying IMCs plasmids. This work was funded by the Department of Biotechnology (DBT), India (Project Code BT/GET/119/SP31652/2020). The Junior Research Fellowship (December 2021–December 2023) and Senior Research Fellowship (December 2023–Present) to H.B. was supported by the Department of Biotechnology (DBT), India.

## Author contributions

A.G. performed the experiments and assay standardizations and finalized the manuscript.

H.B. performed the experiments and assay standardizations, analyzed the data, and finalized the manuscript.

N.M. conceptualized the study, designed the crRNAs, and provided inputs for troubleshooting throughout the study.

S.S. conceptualized the study, prepared the cDNA from the patient sera, and helped with assay troubleshooting.

T.B. helped with assay troubleshooting.

M.K. helped with the statistical analysis.

M.B. provided the viral disease control samples to test the specificity of the assay.

B.K.D. provided the immunological data of the HIV-1-infected infants.

R.L. provided the samples of HIV-1 infected infants and provided patient care and management. Su.S and K.L. conceptualized and designed the study, edited, revised, and finalized the manuscript.

