## Supplementary Table1, Supplementary Figure 1-5 for "A rapid CRISPR/Cas12a-based assay for the detection of HIV-1 Indian Clade-C infections"

### Supplementary Information

| S.No. | Patient ID | Age Group Infant(<2yrs) /Pediatric(2-15yrs) /Adult(<18yrs) | HIV Serology (IgG and IgM) | CD4 Count (Cells/ml) | Viral Load (RNA/ml) |
|---|---|---|---|---|---|
| 1 | AIIMS702 | Infant | Positive | 1234 | 393489 |
| 2 | AIIMS705 | Infant | Positive | 1905 | 445630 |
| 3 | AIIMS707 | Infant | Positive | 164 | 385690 |
| 4 | AIIMS708 | Infant | Positive | 1300 | 410567 |
| 5 | AIIMS712 | Infant | Positive | 1456 | 4477000 |
| 6 | AIIMS713 | Infant | Positive | 2631 | 2000000 |
| 7 | AIIMS714 | Infant | Positive | 1898 | 829000 |
| 8 | AIIMS715 | Infant | Positive | 1663 | 2260000 |
| 9 | AIIMS716 | Infant | Positive | 1498 | 3490000 |
| 10 | AIIMS717 | Infant | Positive | 1289 | 1119000 |
| 11 | AIIMS718 | Infant | Positive | 2156 | 638000 |
| 12 | AIIMS719 | Infant | Positive | 1540 | 117000 |
| 13 | AIIMS721 | Infant | Positive | 1634 | 785000 |
| 14 | AIIMS724 | Infant | Positive | 1540 | 829000 |
| 15 | AIIMS725 | Infant | Positive | 1637 | 2260000 |
| 16 | AIIMS726 | Infant | Positive | 1569 | 3490000 |
| 17 | AIIMS727 | Infant | Positive | 5482 | 1119000 |
| 18 | AIIMS728 | Infant | Positive | 2588 | 638000 |
| 19 | AIIMS729 | Infant | Positive | 2891 | 117000 |
| 20 | AIIMS739 | Infant | Positive | 2289 | 829000 |
| 21 | AIIMS744 | Infant | Positive | 2105 | 117000 |
| 22 | AIIMS747 | Infant | Positive | 2684 | 564720 |
| 23 | AIIMS748 | Infant | Positive | 862 | NA |
| 24 | AIIMS861 | Pediatric | Positive | 360 | NA |
| 25 | AIIMS881 | Adult | Positive | 180 | NA |
| 26 | AIIMS893 | Adult | Positive | NA | NA |
| 27 | AIIMS916 | Infant | Positive | 373 | NA |
| 28 | AIIMS924 | Adult | Positive | 609 | NA |
| 29 | AIIMS967 | Adult | Positive | NA | NA |
| 30 | AIIMS979 | Adult | Positive | NA | NA |
| 31 | AIIMS988 | Infant | Positive | NA | NA |
| 32 | AIIMS078 | Infant | Positive | NA | NA |
| 33 | AIIMS232 | Adult | Positive | 223 | NA |
| 34 | AIIMS261 | Adult | Positive | 49 | NA |
| 35 | AIIMS325 | Adult | Positive | NA | NA |
| 36 | AIIMS326 | Infant | Positive | NA | NA |
| 37 | AIIMS353 | Adult | Positive | 248 | NA |
| 38 | AIIMS598 | Pediatric | Positive | NA | NA |
| 39 | AIIMS619/14 | Infant | Positive | NA | NA |
| 40 | AIIMS619/16 | Pediatric | Positive | NA | NA |
| 41 | AIIMS623 | Adult | Positive | NA | NA |

**Supplementary Table 1: Demographics and immunological profile of the 41 HIV-1 positive donors whose serum samples were used to evaluate the CRISPR/Cas12a-based assay.**

**a**

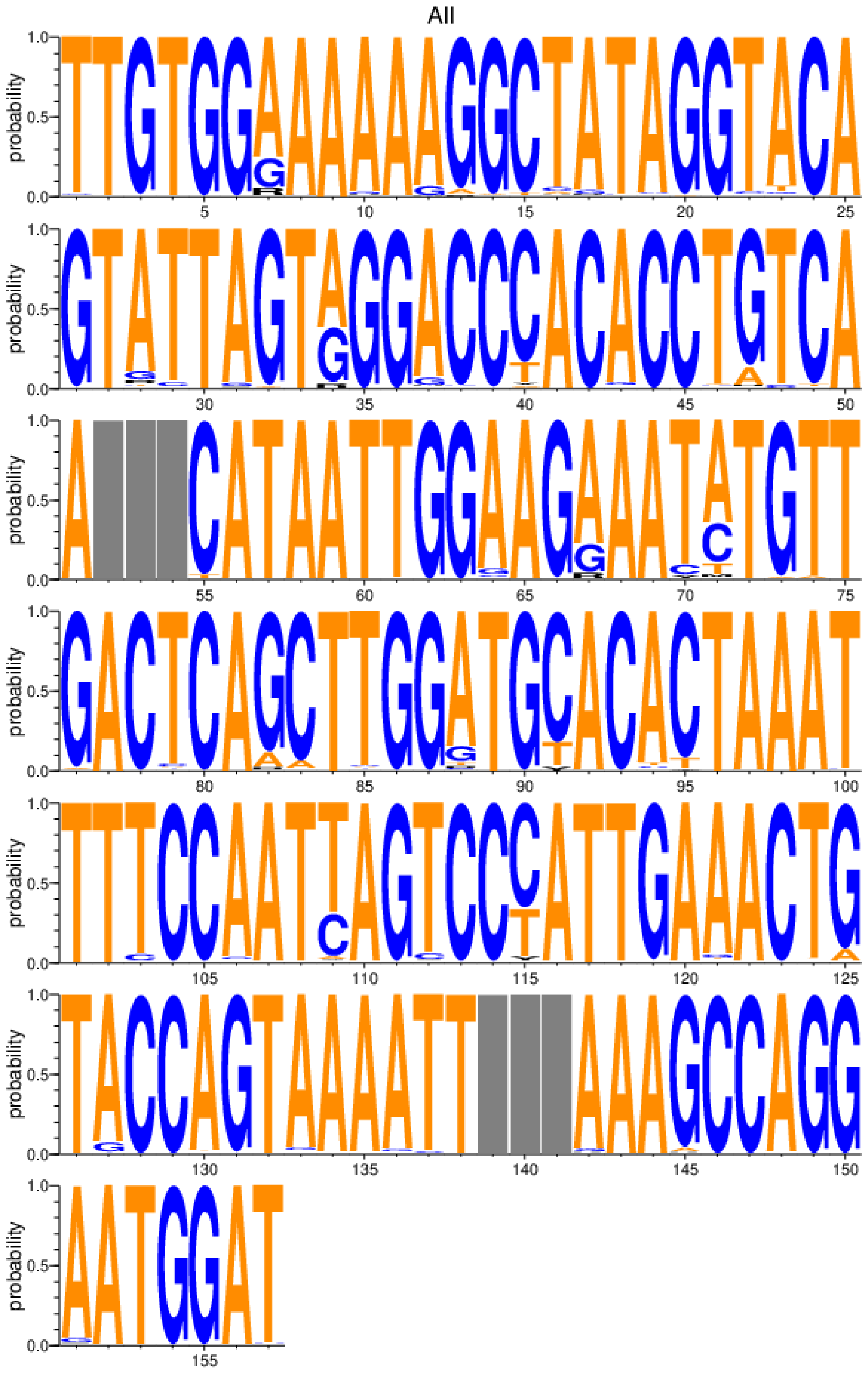

**b**

```
Percentage and raw
count of non-gap  Non-gap/total
(percentage)      Gap/total
(percentage)
1      T: 97.98% (2517)   other: 2.02% (52)      2569/2569 (100.00%)     0/2569 (0.00%)
2      T: 99.69% (2561)   other: 0.31% (8) 2569/2569 (100.00%)     0/2569 (0.00%)
3      G: 99.03% (2543)   other: 0.97% (25)      2568/2569 (99.96%)      1/2569 (0.04%)
4      T: 99.34% (2552)   other: 0.66% (17)      2569/2569 (100.00%)     0/2569 (0.00%)
5      G: 99.96% (2568)   A: 0.04% (1)     2569/2569 (100.00%)     0/2569 (0.00%)
6      G: 99.84% (2565)   other: 0.16% (4) 2569/2569 (100.00%)     0/2569 (0.00%)
7      A: 75.36% (1936)   G: 18.68% (480)   R: 5.37% (138)   other: 0.58% (15) 2569/2569 (100.00%)
8      A: 98.99% (2543)   other: 1.01% (26)      2569/2569 (100.00%)     0/2569 (0.00%)
9      A: 99.84% (2565)   other: 0.16% (4) 2569/2569 (100.00%)     0/2569 (0.00%)
10     A: 96.92% (2490)   other: 3.08% (79)      2569/2569 (100.00%)     0/2569 (0.00%)
11     A: 99.38% (2553)   other: 0.62% (16)      2569/2569 (100.00%)     0/2569 (0.00%)
12     A: 92.64% (2380)   G: 6.50% (167)   other: 0.86% (22) 2569/2569 (100.00%)     0/2569 (0.00%)
13     G: 95.25% (2447)   other: 4.75% (122)     2569/2569 (100.00%)     0/2569 (0.00%)
14     G: 98.52% (2531)   other: 1.48% (38)      2569/2569 (100.00%)     0/2569 (0.00%)
15     C: 97.24% (2498)   other: 2.76% (71)      2569/2569 (100.00%)     0/2569 (0.00%)
16     T: 93.97% (2414)   C: 2.84% (73)   other: 3.19% (82)  2569/2569 (100.00%)     0/2569 (0.00%)
17     A: 95.48% (2453)   other: 4.52% (116)     2569/2569 (100.00%)     0/2569 (0.00%)
18     T: 98.48% (2530)   other: 1.52% (39)      2569/2569 (100.00%)     0/2569 (0.00%)
19     A: 97.47% (2504)   other: 2.53% (65)      2569/2569 (100.00%)     0/2569 (0.00%)
20     G: 98.91% (2541)   other: 1.09% (28)      2569/2569 (100.00%)     0/2569 (0.00%)
21     G: 99.73% (2562)   other: 0.27% (7) 2569/2569 (100.00%)     0/2569 (0.00%)
22     T: 96.46% (2478)   other: 3.54% (91)      2569/2569 (100.00%)     0/2569 (0.00%)
23     A: 93.11% (2392)   T: 3.93% (101)   other: 2.96% (76) 2569/2569 (100.00%)     0/2569 (0.00%)
24     C: 99.96% (2568)   S: 0.04% (1)     2569/2569 (100.00%)     0/2569 (0.00%)
25     A: 99.10% (2546)   other: 0.90% (23)      2569/2569 (100.00%)     0/2569 (0.00%)
26     G: 99.49% (2556)   other: 0.51% (13)      2569/2569 (100.00%)     0/2569 (0.00%)
27     T: 99.84% (2565)   other: 0.16% (4) 2569/2569 (100.00%)     0/2569 (0.00%)
28     A: 89.65% (2303)   G: 5.29% (136)   R: 2.34% (60)   other: 2.72% (70)   2569/2569 (100.00%)
29     T: 95.68% (2458)   other: 4.32% (111)     2569/2569 (100.00%)     0/2569 (0.00%)
30     T: 99.96% (2568)   C: 0.04% (1)     2569/2569 (100.00%)     0/2569 (0.00%)
31     A: 96.46% (2478)   other: 3.54% (91)      2569/2569 (100.00%)     0/2569 (0.00%)
32     G: 98.72% (2536)   other: 1.28% (33)      2569/2569 (100.00%)     0/2569 (0.00%)
```

**Supplementary Figure 1: Identification of conserved regions in HIV-1 Clade-C sequences for designing of crRNA a)** Conserved regions in the Pol gene obtained by alignment of 86-HIV-1 Clade C sequences queried through the HIV sequence database (https://www.hiv.lanl.gov/content/sequence/HIV/mainpage.html) for the design of crRNA design. **b)** Frequency table of the aligned of the HIV-1 Clade C sequences for identification of the conserved region in Pol region for the design of crRNA.

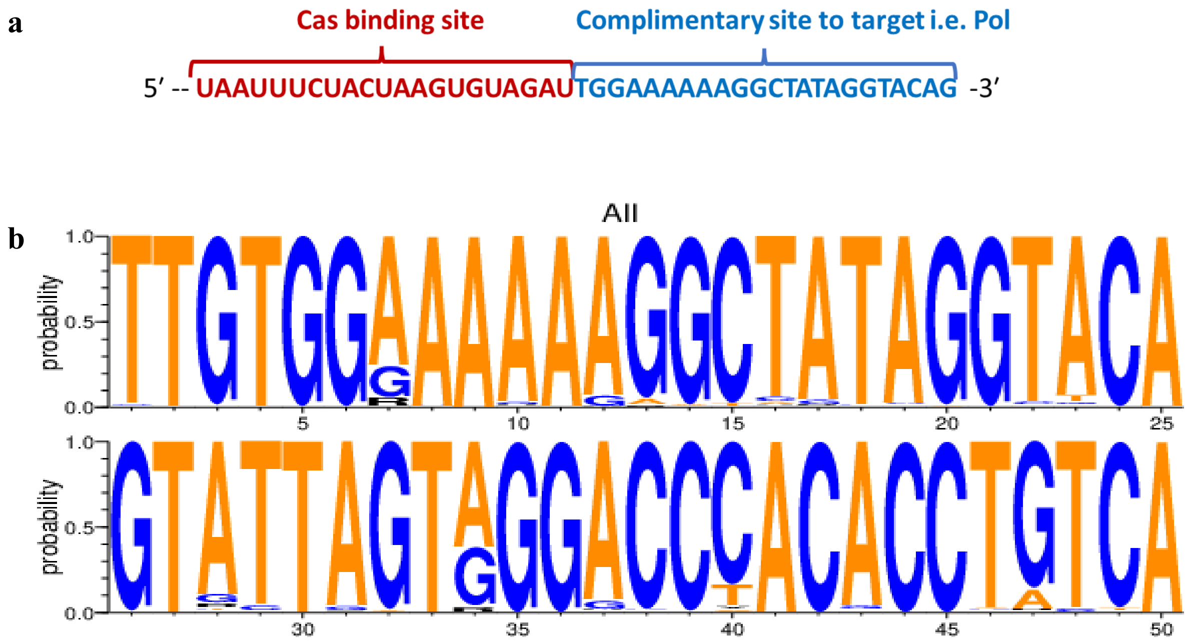


**Supplementary Figure 2: Characteristics of the crRNA designed against the conserved region of HIV-1 Pol gene of Indian Clade-C sequences. a)** Nucleic acid sequence of the crRNA. Sequences in blue represent the complementary region in the HIV-1 Pol gene, and the sequences in red represent the Cas12a binding site. **b)** The sequence logo plot depicts the frequency of the crRNA sequence bases and flanking region in the Pol region against which the crRNA was designed.

**a** Primer for RPA:

- RPA_Forward - 5’-ACAGGRGCAGATGATACAGTATTAGAAGA-3’
- RPA_Reverse - 5’-CCAATTATGTTGAYAGGTGTDGGTCC-3’

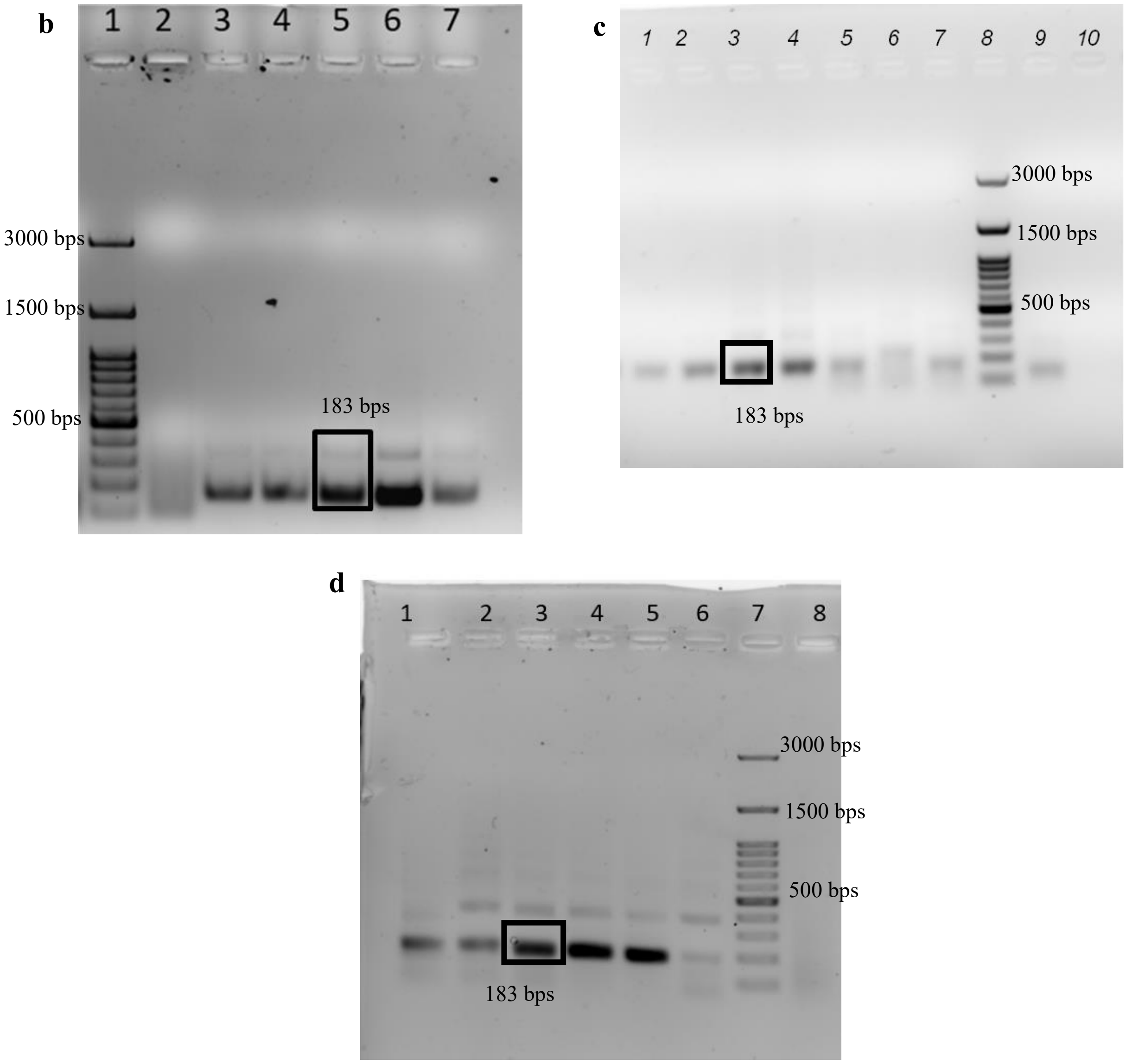


**Supplementary Figure 3: RPA standardization. a)** Degenerate primer pairs for the RPA amplicon. **b)** 1.5% Agarose gel run for RPA time gradient with positive control(93IN999): 1st lane – ladder(100bp), 2nd lane - NTC, 3rd lane -20min, 4th lane -25min, 5th lane -30min, 6th lane -35min,7th lane -40min. **c)** 1.5% Agarose gel run for RPA primer gradient with positive control(93IN999): lane 1 -144nM, 2nd lane - 156nM, 3rd lane -168nM, 4th lane -180nM, 5th lane -192nM, 6th lane -204nM,7th lane -216nM,8th lane – ladder(100bp),9th lane -228nM,10th lane -NTC. **d)** 1.5% Agarose gel run for RPA standardization of patient cDNA (AIIMS748): 1st lane-AIIMS748 neat cDNA, 2nd lane- AIIMS748 (1:5) dilution, 3rd lane-AIIMS748(1:10) dilution, 4th lane- 93IN999, 5th lane- 93IN905, 6th lane- AIIMS748(1:20) dilution, 7th lane- ladder(100bp),8th lane- NTC.

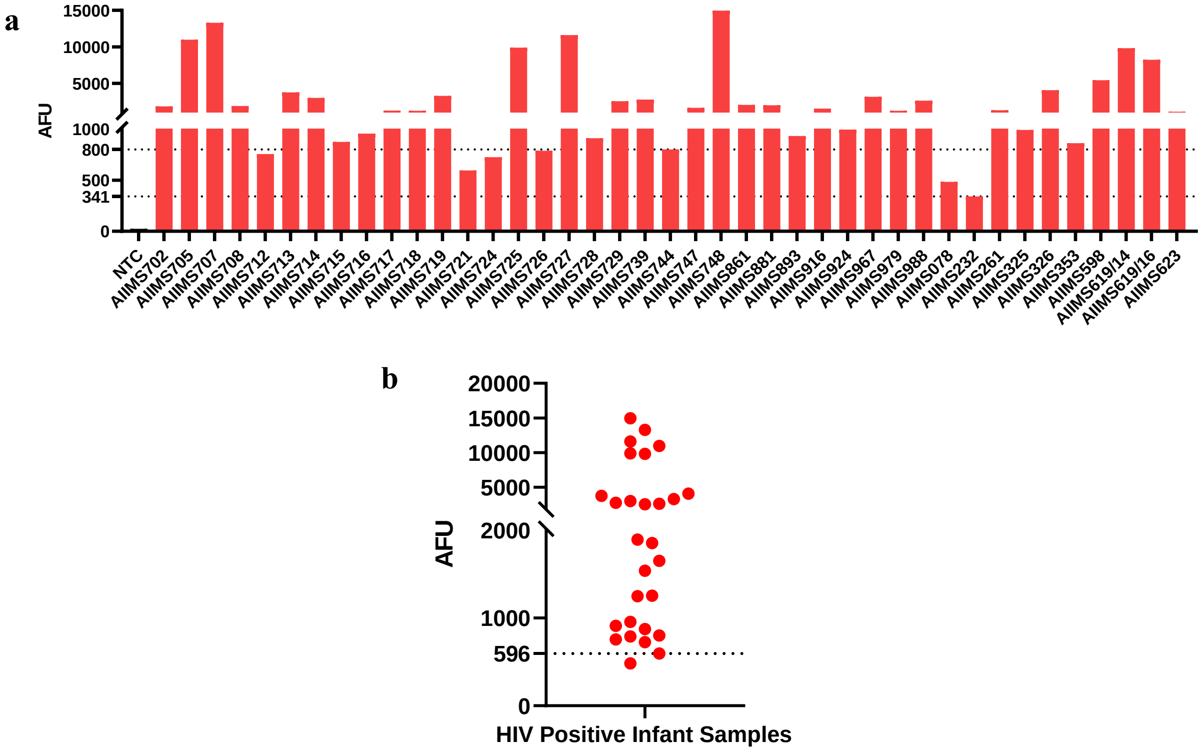


**Supplementary Figure 4: Validation of the CRISPR/Cas12a-based assay for detecting HIV-1 Clade-C nucleic acid in positive clinical samples. a)** Fluorescence readout of the assay performed on 41 HIV-1 positive samples. The fluorometer readings of fluorescence emission were recorded as AFU (Arbitrary fluorescence unit) at excitation at 488nm and emission at 520nm. AFU threshold values 800 (highest) and 341(lowest) were used to distinguish positive and negative results of the assay. **b)** Fluorescence readout of the assay performed on 28 HIV-1 positive infant samples. AFU threshold value of 596 was used to distinguish the positive and negative results of the assay.

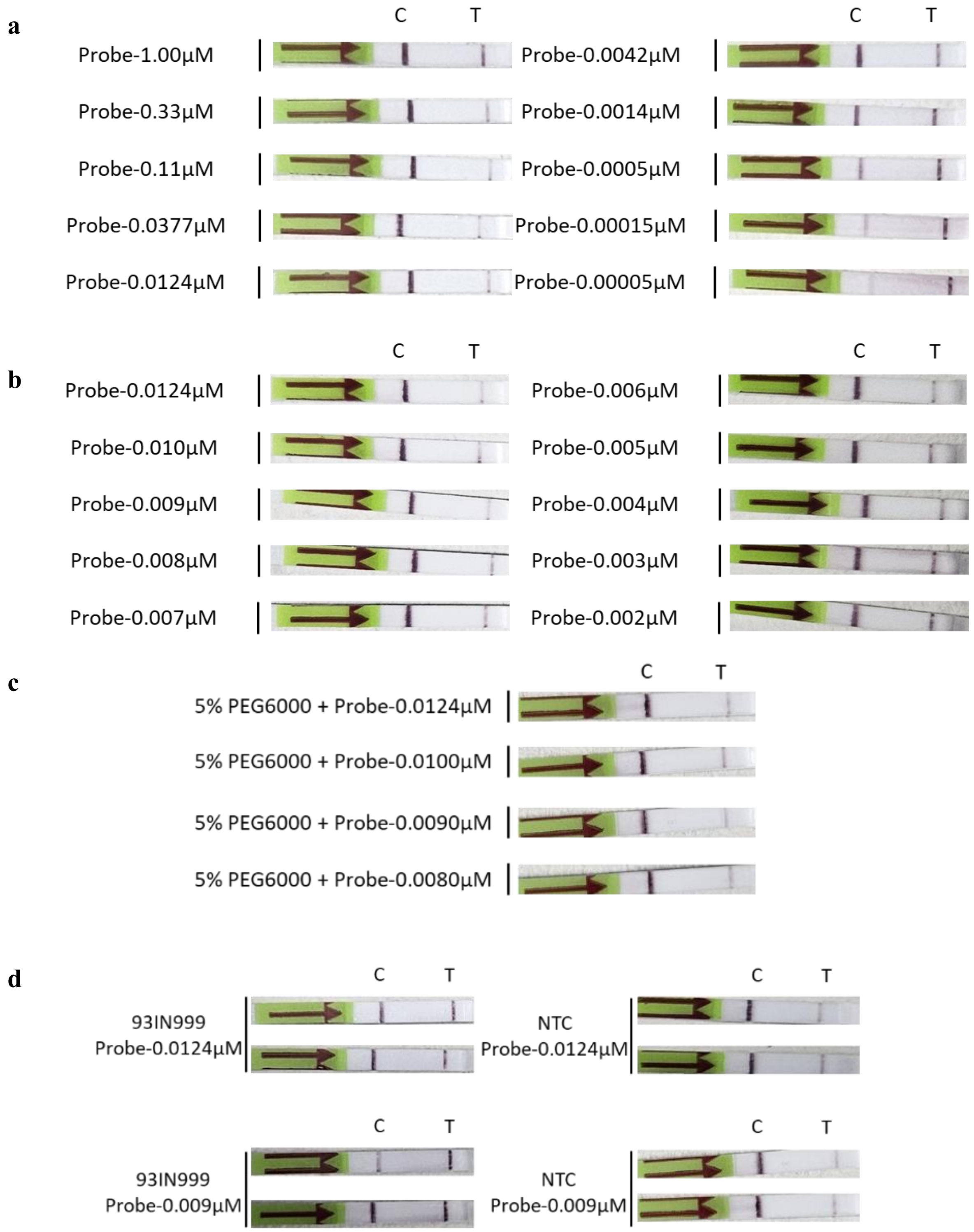


**Supplementary Figure 5: Standardization of CRISPR/Cas12a for lateral flow strip-based detection of HIV-1 Clade C nucleic acid. a)** Optimization of ssDNA-FAM-Biotin probe concentration for minimal non-specific detection on the test line in NTCs (HIV-1 genome plasmid not added) for a range of probe concentrations (broad range): 1.00µM to 0.00005µM. **b)** Determination of optimal probe concentration using the concentrations between 0.0124µM and 0.002µM (narrow range). **c)** Increasing the specificity of lateral flow-based detection by minimizing the detection of the probe at the test band with the addition of PEG6000 for a range of probe concentrations: 0.0124μM to 0.008μM. **d)** Detection of CRISPR/Cas12a-based assay with lateral flow strips using HIV-1 genome plasmid-93IN999 using two of the most optimal probe concentrations: 0.0124μM and 0.009μM. A dark band on the test line represents a positive detection with no detection on the test line in the corresponding NTC strip.
